# Pattern and distribution of cancers in areas of Iraq exposed to Depleted Uranium

**DOI:** 10.1101/2020.04.10.20060475

**Authors:** Esraa Aldujaily, Ali Duabil, Kussay M. Abbas Zwain, Hayder K. Fatlawi, Ali Al-Behadili, Emad Al saabery, Yasseen A. Alwaaely, Sijal Al Joboury, Omar Layth Qassid, Liwaa Al Kelabi

## Abstract

**Introduction:** Cancer is one of the major causes of death worldwide. Health systems whether in developed or in developing countries like Iraq are burdened with different programs to control cancer. Our study is intended to provide information about cancer in the region of Middle Euphrates Area (MEA) of Iraq, which is one of the major areas in Iraq that exposed to the depleted Uranium (DU) at different time periods. Therefore, we are aiming to explore more information about the behavior of cancers in this region of Iraq (pattern and distribution).

Aim: our study aims to describe the landscape of cancer with wide focus on the clinicopathological behavior of different types of cancers in MEA of Iraq to determine whether any differences have cropped up over time in Iraqi patients’ presentations.

**Patients and methods:** This study is a retrospective descriptive study design. Data were collected from a single tertiary cancer care oncology centre for three consecutive years from 2016 up to 2018. This Database covers nearly the entire Middle Euphrates area of Iraq. All statistical tests performed at a 95% level of significance with a two-sided p-value of 0.05 indicating statistical significance.

**Results and conclusion:** According to this study, the three most common cancers among the entire population were breast, lung, and brain cancers. Females constituted 57.0% of the entire study. Most cancers including breast cancer presented with aggressive clinicopathological behavior. Middle age groups of both sexes are more at risk of developing different cancers. Such findings are important and pave the way for future scientific cancer control programs in Iraq especially for breast cancer. The cancer appears to be flourishing in Iraq, which could be due to multiple factors. Finding a new strategy to predict the treatment response, recurrence or aggressiveness of cancers in Iraq is crucial.

**Summary (Strengths and limitations of this study):** Whats new in this study is the wide focus of studying the clinicopathological behavior of all cancers in middle Euphrates area of Iraq to establish a solid base for further future studies that aims at developing the health system and cancer control programs in Iraq and middle east. The area of middle and south of Iraq which is named the middle Euphrates area is an area of major interest for researchers and clinicians as the cancer is flourishing in this area. previous studies excuse this increase of cancer incidence to the exposure to cancerous agents like depleted uranium. This fact inspired us to explore more information about the behavior of cancer in Iraq. The limitations of this study are the lack of therapy and survival data that we hope to include them in future studies.

## Introduction

Globally, cancers have become a leading cause of morbidity and mortality, placing significant burden on health systems and the society at large, especially in developing countries and developed countries. Latest global data on cancer from the International Agency for Research on Cancer (IARC) indicates that there were as much as 18.1 million new cancer cases and 9.6 million cancer-related deaths in 2018.^1^ The burden is such that at least one in every five men, and one in every six women is expected to develop the disease during their lifetime.^1^ In Iraq, GLOBOCAN reported 25,320 new cases of cancers in 2018, with 14,020 (55.37%) in women and 11,300 (44.62%) in men.^2^ With the number of deaths caused by cancers, it is unsurprising that cancers are now regarded as the second cause of mortalities globally.^3,4^ It is noteworthy that the cancer mortality rate in Iraq is comparable with results from many developing countries, as 57.36% of the new cases in 2018 died as a result of their disease.^2^ It is poor cancer indices such as these that have led to increasing attention on cancer control in Iraq.

Over the years, there has been a lot of interest in understanding the reasons for the increasing burden of cancer in Iraq and surrounding nations. Evidence from existing literature suggest that the rising cancer incidence rate is as a result of the ageing population; shift to cancer-promoting lifestyles including smoking, excessive alcohol consumption, physical inactivity; obesity, lower parity, delayed first birth; all of which are associated with improving economic development, urbanization, and westernization.^5,6^ Other authors have pointed at the increasing awareness, availability of cancer diagnostic centers, and better registration of suspected cancer cases as contributing to the rising numbers of cancer patients in the region.^7,8^ Another unique factor which has also been implicated in the changing face of cancer in Iraq is the extensive damage to the environment as a result of war, exposure to depleted Uranium, and associated economic sanctions.^9,10^

There have been significant efforts in Iraq to control the increasing incidence of cancers, especially with the provision of cancer health services, launching of public health campaigns to encourage early detection of cancers, encouraging physical activity and control of tobacco use, and providing better access to cancer diagnosis and treatment service free of charge. Recent data from the UICC indicates that the establishment of the Iraq population-based cancer registry as well as cancer diagnosis and treatment centers in Baghdad and other provinces have led to significant progress towards cancer control in Iraq.^7,8^ These have particularly led to increased awareness, early cancer detection and treatment, and improved access to cancer care in the country.

However, there are still huge gaps in the efforts to control the rising cancer incidence in Iraq. The collection of data via the country’s cancer registry and conduct of a number of site-based cancer epidemiological studies have attempted to shed more light on the pattern of cancers the country. Studies conducted in country have shown that disease presentation is often affected by the ethnic variation of the population.^7,11–13^ Data from the national registry which would have been a great resource for understanding the ethnic differences in cancer presentation is often incomplete as it is deficient of details such as clinical and pathological profile of cancer patients suggesting a need for studies describing the pattern of presentation of patients with cancers.^14,15^ With increasing interest in developing effective methods of cancer control and patient-oriented cancer care programs in Iraq, it becomes even more important to conduct deep-dive studies providing detailed information on the sociodemographic, clinical, and pathologic profiles of Iraqi cancer patients.

This study aims at adopting scientific approach to the clinicopathological parameters of cancer patients in Iraq to support the efforts for developing the strategies for cancer control in Iraq by providing detailed information about the pattern, distribution, and clinical characteristics of cancers in the Middle Euphrates region of the country. This region of Iraq exposed to depleted Uranium at different time periods during wars and conflicts.^9^

Specifically, the objectives of the study are to (i) determine the patterns of presentation of cancer patients over a 3-year period, and (ii) describe the time trend of the pattern of distribution of breast cancers in Middle Euphrates area using an oncology database. The results of this study will provide a crucial baseline for future cancer epidemiological and genetic studies, as well as an input into the development of Artificial model (AI) models to predict treatment response, recurrence, or aggressiveness; and guide cancer control efforts in Iraq and the entire Middle Eastern region.

## Methodology

### Study setting

The study was conducted at Middle Euphrates Oncology centre in Najaf which is a 100-bed tertiary cancer care center that primarily receives in-patients and outpatients from Najaf city and functions as a referral center for other cities in the entire Middle Euphrates area. The facility uniquely has a detailed cancer database that houses information for patients managed for a wide variety of cancers.

### Patient and public involvement

No patient involved.

### Study design

The study utilized a retrospective descriptive study design to determine the pattern, distribution, and clinical characteristics of cancers in patients presenting for treatment between 2016 and 2018 at the Middle Euphrates Oncology Centre in Najaf city, Iraq. All patients seen and managed for all cases of cancers between January 2016 and December 2018 were eligible for inclusion in this study. Data was collected only on patients of all sexes with all types of cancer distributed by grade, metastasis, recurrence, sex, site, invasion, stage and stage group as confirmed by biopsy, blood work, computed tomography (CT), cytology, histology, and/or magnetic resonance imaging (MRI). Patients with mental deficits precluding proper follow-up and documentation and those with incomplete information were excluded from the study.

### Data collection and analysis

Data was extracted from the oncology database using a proforma developed for the purpose. Members of the research team received training to ensure consistency and fidelity during the process of data collection. Sociodemographic, clinical, and histopathological variables were extracted. Data on sociodemographic variables such as age and gender of the patients was collected. Clinical variables included cancer site, tumor size (for solid cancers), disease stage, possibility of recurrence, and presence of distant metastases. Histopathological variables included cell grade, invasion factor, and the histological type of the malignancy. It is important to note that some of the data in the database was collected directly from patients, and in a few instances, from first-degree relatives serving as caregivers.

The primary objective of this study was to determine the patterns of the top 10 cancers over a 3-year period with a secondary focus on breast cancers. Using the R program for statistical analysis, descriptive statistics were carried out and summarized in the form of means and standard deviation (SD), for continuous data, while frequencies and percentages were used for categorical data. Univariate tests (chi square) and the student’s t-test were carried out on categorical and continuous variables respectively. All statistical tests performed at a 95% level of significance with a two-sided p-value of 0.05 indicating statistical significance.

### Ethical implications

Approval to carry out the study was granted by the Iraqi Ministry of Higher Education and Scientific Research Ethics Committee (Medical Ethics Committee, MEC-20); and study conduct followed the tenets of the Helsinki Declaration. Each patient’s data was covered by full privacy policies, and each patient’s identity and profile were fully anonymized and protected.

## Results

### Demographic distribution

A total of 4,852 patients were seen and managed for different types of cancers between 2016 and 2018 at the oncology center, with 1,891 seen in 2016, 1,360 in 2017, and 1,601 in 2018. Females constituted 57.0% of the entire study population, and the mean age of all the patients seen across the three years varied between 50.99 and 52.77 years (Table 1).

**Table 1.** Demographic characteristics of the study population

| Variable | Years |  |  | Total |
| --- | --- | --- | --- | --- |
|  | 2016 (n = 1,891) | 2017 (n = 1,360) | 2018 (n = 1,601) |  |
| Sex |  |  |  |  |
| Female | 1,025 (54.41%) | 809 (59.79%) | 923 (57.80%) | 2757 (57.03%) |
| Male | 859 (45.59%) | 544 (40.21%) | 674 (42.20%) | 2077 (42.97%) |
| **Age** |  |  |  |  |
| **Mean (SD)** | 51.74 (18.35%) | 50.99 (18.69%) | 52.77 (17.82%) | NA |

### Pattern of presentation – clinical profile

Primary cancers were found in a total of 4,796 patients, constituting 97.90% of the entire population; and secondary cancers in the remaining 1.15%. As seen in Fig. 1, the ten most common types of cancers in males were lung, bladder, brain, prostate, non-Hodgkin’s lymphoma (NHL), leukemia, larynx, soft tissue, Hodgkin’s disease, and bone cancers. In females, the first ten were breast, lung, brain, uterine, ovarian, NHL, soft tissue, urinary bladder, Hodgkin’s disease, and cervical cancers.

**Figure 1.**
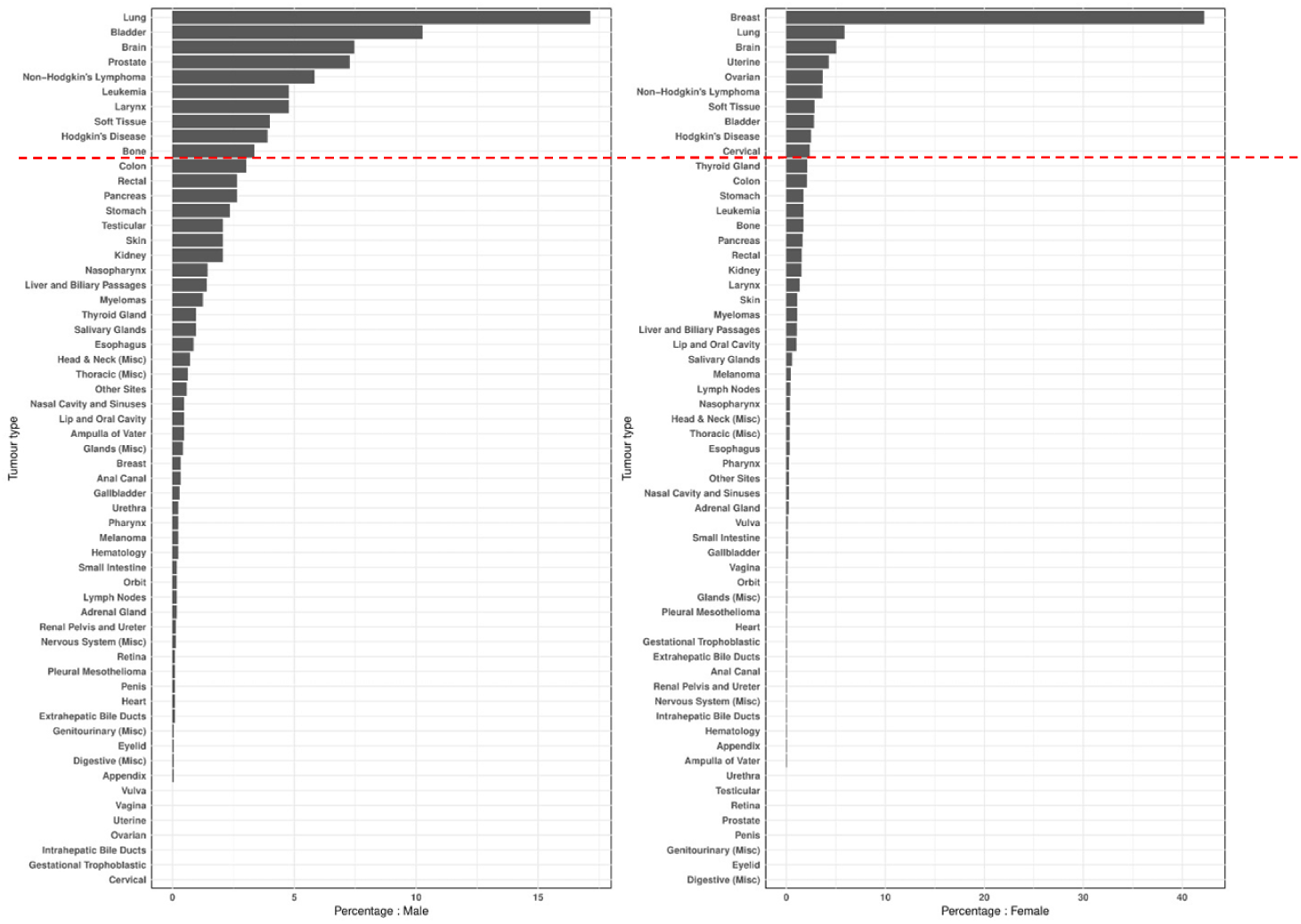
Distribution of cancers by site in males and females *(red line indicating the top ten cancers in each gender category)*

Going further to examine the disease type by primary or secondary cases, most of the patients had primary disease (Table 2). Across the three years, 2016 – 2018, majority of the patients presented with disease in Table 3 indicates that majority of the patients presented with Grade II disease (21.35%), followed by those with Grade III and Grade I with proportions of 14.41% and 11.31% respectively. Furthermore, of the cases with known recurrence status, only a small proportion had recurrent disease ranging from 3.44% in 2016 to 3.60% in 2017 and 1.25% in 2018. The proportion of patients who had distant metastases grew over the years from 18.51% to 23.61% between 2016 and 2018.

**Table 2.** Clinical profile of the study population

| Variable | Years (n, %) |  |  | Total |
| --- | --- | --- | --- | --- |
|  | 2016 | 2017 | 2018 |  |
| Ranking |  |  |  |  |
| Primary | 1,866 (98.68%) | 1,346 (98.97%) | 1,584 (98.94%) | 98.85% |
| Secondary | 25 (1.32%) | 14 (1.03%) | 17 (1.06%) | 1.15% |
| Recurrence |  |  |  |  |
| Not Recurrence | 720 (38.08%) | 1,254 (92.21%) | 1,572 (98.19%) | 73.08% |
| Recurrence | 65 (3.44%) | 49 (3.60%) | 20 (1.25%) | 2.76% |
| Unknown | 1,106 (58.49%) | 57 (4.19%) | 9 (0.56%) | 24.15% |
| **Metastasis** |  |  |  |  |
| Distant metastasis | 350 (18.51%) | 285 (20.96%) | 378 (23.61%) | 20.88% |
| No Distant metastasis | 319 (16.87%) | 641 (47.13%) | 907 (56.65%) | 38.48% |
| Unknown | 1,222 (64.62%) | 434 (31.91%) | 316 (19.74%) | 40.64% |
| **Cell Grade** |  |  |  |  |
| Grade I | 196 (10.36%) | 156 (11.47%) | 197 (12.30%) | 11.31% |
| Grade II | 398 (21.05%) | 289 (21.25%) | 349 (21.80%) | 21.35% |
| Grade III | 236 (12.48%) | 239 (17.57%) | 224 (13.99%) | 14.41% |
| Grade IV | 89 (4.71%) | 28 (2.06%) | 45 (2.81%) | 3.34% |
| Grade X | 498 (26.34%) | 513 (37.72%) | 275 (17.18%) | 26.50% |
| Unknown | 474 (25.07%) | 135 (9.93%) | 511 (31.92%) | 23.08% |

**Table 3.** Histopathological profile of the study population

| Variables | Years (n, %) |  |  | Total |
| --- | --- | --- | --- | --- |
|  | 2016 | 2017 | 2018 |  |
| Invasive |  |  |  |  |
| Micro-invasive | 4 (0.21%) | 0 (0.00%) | 1 (0.06%) | 5 (0.10%) |
| Non-invasive | 14 (0.74%) | 9 (0.66%) | 10 (0.62%) | 33 (0.68%) |
| Fully invasive | 156 (8.25%) | 37 (2.72%) | 14 (0.87%) | 207 (4.27%) |
| Unknown | 1717 (90.70%) | 1314 (96.64%) | 1576 (98.44%) | 4607 (94.95%) |

| Stage |  |  |  |  |
| --- | --- | --- | --- | --- |
| Accelerated | 0 (0.00%) | 1 (0.07%) | 0 (0.00%) | 1 (0.02%) |
| Blast | 6 (0.32%) | 20 (1.47%) | 26 (1.62%) | 52 (1.07%) |
| Chronic | 2 (0.11%) | 2 (0.15%) | 1 (0.06%) | 5 (0.10%) |
| Occult | 0 (0.00%) | 0 (0.00%) | 1 (0.06%) | 1 (0.02%) |
| Stage 0 | 0 (0.00%) | 2 (0.15%) | 4 (0.25%) | 6 (0.12%) |
| Stage 0a | 2 (0.11%) | 0 (0.00%) | 1 (0.06%) | 3 (0.06%) |
| Stage 0is | 0 (0.00%) | 1 (0.07%) | 0 (0.00%) | 1 (0.02%) |
| Stage I | 18 (0.95%) | 22 (1.62%) | 46 (2.87%) | 86 (1.77%) |
| Stage IA | 7 (0.37%) | 22 (1.62%) | 33 (2.06%) | 62 (1.28%) |
| Stage IB | 4 (0.21%) | 9 (0.66%) | 18 (1.12%) | 31 (0.64%) |
| Stage IB1 | 0 (0.00%) | 1 (0.07%) | 0 (0.00%) | 1 (0.02%) |
| Stage IC | 0 (0.00%) | 1 (0.07%) | 3 (0.19%) | 4 (0.08%) |
| Stage II | 11 (0.58%) | 22 (1.62%) | 29 (1.81%) | 62 (1.28%) |
| Stage IIA | 33 (1.75%) | 59 (4.34%) | 70 (4.37%) | 162 (3.34%) |
| Stage IIB | 29 (1.53%) | 91 (6.69%) | 78 (4.87%) | 198 (4.08%) |
| Stage IIC | 0 (0.00%) | 0 (0.00%) | 1 (0.06%) | 1 (0.02%) |
| Stage III | 10 (0.53%) | 17 (1.25%) | 24 (1.50%) | 51 (1.05%) |
| Stage IIIA | 37 (1.96%) | 78 (5.74%) | 95 (5.93%) | 210 (4.33%) |
| Stage IIIB | 13 (0.69%) | 30 (2.21%) | 57 (3.56%) | 100 (2.06%) |
| Stage IIIC | 19 (1.00%) | 32 (2.35%) | 41 (2.56%) | 92 (1.90%) |
| Stage IIIC1 | 0 (0.00%) | 0 (0.00%) | 1 (0.06%) | 1 (0.02%) |
| Stage IS | 0 (0.00%) | 0 (0.00%) | 1 (0.06%) | 1 (0.02%) |
| Stage IV | 104 (5.50%) | 142 (10.44%) | 286 (17.86%) | 532 (10.96%) |
| Stage IVA | 6 (0.32%) | 6 (0.44%) | 22 (1.37%) | 34 (0.70%) |
| Stage IVB | 26 (1.37%) | 35 (2.57%) | 48 (3.00%) | 109 (2.25%) |
| Stage IVC | 4 (0.21%) | 7 (0.51%) | 11 (0.69%) | 22 (0.45%) |
| Stage X | 484 (25.59%) | 426 (31.32%) | 310 (19.36%) | 1220 (25.14%) |
| Unknown | 1,076 (56.90%) | 334 (24.56%) | 394 (24.56%) | 1804 (37.18%) |

### Histopathological profile of the study population

On histology, the invasiveness status of many of the cancer patients was not stated. However, for those that had data on the presence of invasive disease, 207 (4.27%) had invasive disease (Table 3). Only 5 patients were reported to have micro-invasive disease. Similar to the grade distribution of the patients, majority were in stage IV (n = 532, 10.96%) at presentation, followed by those who were in stage IIIA (n = 210, 4.33%) and stage IIB (n = 198, 4.08%).

### Breast cancer profile

A deep dive into the 1,164 patients with breast cancers showed that almost 100% of them were unilateral primary cases with 50.8% of the patients with known laterality having their cancers on the left side, and 48.8% on the right side (Table 4). A very small minority had bilateral disease (0.3%). The mean ages of the patients varied from 49.4 years to 50.8 years across the three years of study. Pathologically, the most predominant form of breast cancer seen was the ductal type, with 965 (82.90%) patients suffering from this. The next two most common pathological forms of the disease were the lobular breast cancer (99, 8.51%) and carcinoma (37, 3.18%). Majority of the patients presented with stage IIIA disease (146, 12.54%), followed by those in stage IIB (134, 11.51%) and stage IIA (121, 10.40%).

**Table 4.** Demographic, clinical, and pathological profile of patients with breast cancer (n = 1164)

| Variables | Years (n, %) |  |  | Total |
| --- | --- | --- | --- | --- |
|  | 2016 | 2017 | 2018 |  |
| Age |  |  |  |  |
| Mean (SD), *years* | 49.35 (11.47) | 50.76 (11.64) | 50.61 (11.74) | NA |
| Minimum | 21 | 20 | 24 | NA |
| Maximum | 91 | 83 | 91 | NA |
| Ranking |  |  |  |  |
| Primary | 420 (99.53%) | 375 (100.00%) | 367 (100.00%) | 1162 (99.83%) |
| Secondary | 2 (0.47%) | 0 (0.00%) | 0 (0.00%) | 2 (0.17%) |
| Laterality |  |  |  |  |
| Both sides | 0 (0.00%) | 2 (0.53%) | 2 (0.54%) | 4 (0.34%) |
| Left side | 47 (11.14%) | 169 (45.07%) | 171 (46.59%) | 387 (33.25%) |
| Right side | 55 (13.03%) | 167 (44.53%) | 150 (40.87%) | 372 (31.96%) |
| Unknown | 320 (75.83%) | 37 (9.87%) | 43 (11.72%) | 400 (34.36%) |
| Tumour category |  |  |  |  |
| Adenocarcinoma | 0 (0.00%) | 2 (0.53%) | 2 (0.54%) | 4 (0.34%) |
| Adenoid Cystic | 1 (0.24%) | 0 (0.00%) | 0 (0.00%) | 1 (0.09%) |
| Carcinoma | 6 (1.42%) | 8 (2.13%) | 23 (6.27%) | 37 (3.18%) |
| Carcinosarcoma | 0 (0.00%) | 1 (0.27%) | 0 (0.00%) | 1 (0.09%) |
| Ductal | 353 (83.65%) | 317 (84.53%) | 295 (80.38%) | 965 (82.90%) |
| Fibrosarcoma | 0 (0.00%) | 1 (0.27%) | 0 (0.00%) | 1 (0.09%) |
| Lobular | 22 (5.21%) | 35 (9.33%) | 42 (11.44%) | 99 (8.51%) |
| Medullary | 5 (1.18%) | 5 (1.33%) | 1 (0.27%) | 11 (0.95%) |
| Mucinous | 3 (0.71%) | 1 (0.27%) | 3 (0.82%) | 7 (0.60%) |
| Papillary Adenocarcinoma | 3 (0.71%) | 0 (0.00%) | 0 (0.00%) | 3 (0.26%) |
| Squamous Cell | 0 (0.00%) | 0 (0.00%) | 1 (0.27%) | 1 (0.09%) |
| Tubular Adenocarcinoma | 2 (0.47%) | 0 (0.00%) | 0 (0.00%) | 2 (0.17%) |
| Unknown | 27 (6.40%) | 5 (1.33%) | 0 (0.00%) | 32 (2.75%) |
| **Stage** |  |  |  |  |
| Stage 0 | 0 (0.00%) | 2 (0.53) | 3 (0.82%) | 5 (0.43%) |
| Stage IA | 6 (1.42%) | 14 (3.73%) | 20 (5.45%) | 40 (3.44%) |
| Stage IIA | 28 (6.64%) | 39 (10.40%) | 54 (14.71%) | 121 (10.40%) |
| Stage IIB | 21 (4.98%) | 63 (16.80%) | 50 (13.62%) | 134 (11.51%) |
| Stage IIIA | 29 (6.87%) | 60 (16.00%) | 57 (15.53%) | 146 (12.54%) |
| Stage IIIB | 5 (1.18%) | 9 (2.40%) | 12 (3.27%) | 26 (2.23%) |
| Stage IIIC | 17 (4.03%) | 30 (8.00%) | 32 (8.72%) | 79 (6.79%) |
| Stage IV | 17 (4.03%) | 25 (6.67%) | 57 (15.53%) | 99 (8.51%) |
| Stage X | 174 (41.23%) | 93 (24.80%) | 54 (14.71%) | 321 (27.58%) |
| Unknown | 125 (29.62%) | 40 (10.67%) | 28 (7.63%) | 193 (16.58%) |

As depicted in Figure 2, the receptor classification of the breast cancer patients varied widely between the three receptor types – Human epidermal growth factor receptor-2 (Her2), progesterone receptor (PR), and estrogen receptor (ER). Of the patients with known receptor classification, 276 (61.74%) did not have the Her2 receptor amplified (0 – 1+). Another 38 (8.50%) had equivocal Her2 receptor status while the remaining 133 patients (29.75%) had amplified Her2 receptors. With respect to progesterone receptor status, majority were PR positive (323 patients, 71.62%) while the rest were negative. Similarly, a larger percentage of those with known ER status were ER positive (347, 76.43%) with the remainder being ER negative.

**Figure 2.**
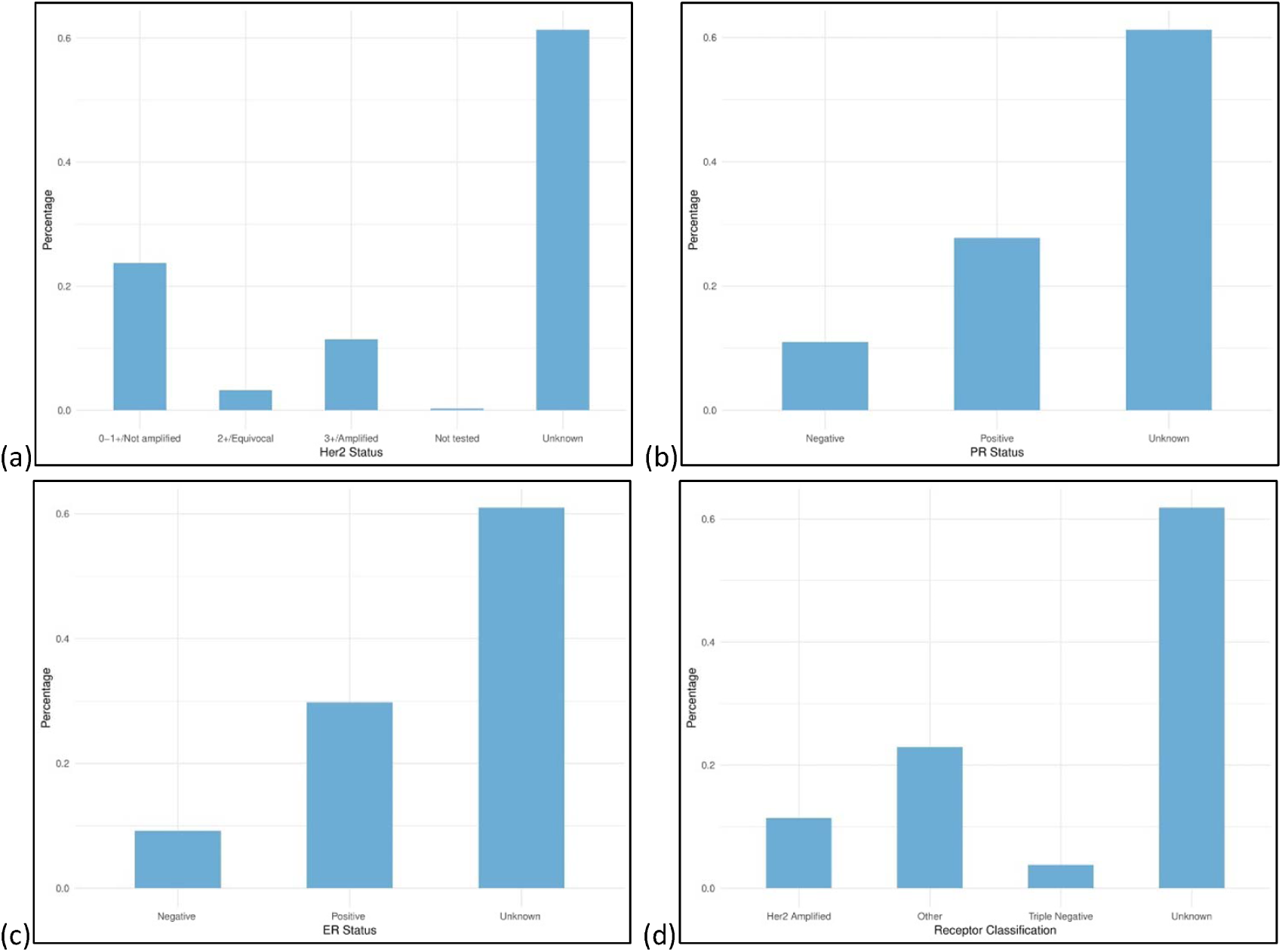
(a) Her2 status, (b) PR status, (c) ER status, and (d) receptor classification of the study population with breast cancer

## Discussion

The results of this study give an overview of the demographic, clinical, and histopathological profile of patients with cancers in Middle Euphrates area, Iraq. The numbers of patients seen for cancers decreased between 2016 and 2018 at the study center, contrasting with previous studies and the general concerns about the increasing incidence of cancers in Iraq^7,14,16,17^ and the rest of the world.^1,4,18^ In 2016, a total of 1,891 cancer patients were seen, dropping sharply to 1,360 in 2017, and rising to 1,601 in 2018. These three data points may not be enough to make a definite conclusion about the cancer presentation, and perhaps, incidence trends in Middle Euphrates area; and may require reviewing the trends over a few more years to decide if this decline in cancer presentations was spurious or erratic.

A female preponderance of cancers was observed in this study, with females constituting almost 57% of all the cases; agreeing with GLOBOCAN cancer incidence estimates of 2018 where 14,020 out of the total of 25,320 new cases were in females.^2^ The mean ages of both sexes put together increased from 51.74 years in 2016 to 52.77 years in 2018. This may not be a major increase, but it aligns with hypotheses that an ageing population may be associated with increases in cancer incidence rates.^5,6^ Also, it suggests a need for increased focus and attention on people in the middle-aged category, particularly with higher index of suspicion for cancers in people 40 years and above. Population cancer screening efforts may need to be targeted at middle aged individuals as a way of picking up the disease, if present, early enough and commence relevant treatment.

A closer look at the clinical profile of the patients studied indicates that the three most common cancers among the entire population were breast, lung, and brain cancers. The first position was driven by one major factor: gender. First, there is the fact that about 57% of the entire population were females, and secondly, there was a less even spread of cancers by location among women, as opposed to men. Figure 1 illustrates this clearly, with male cancers having a gradual slope from lung cancer to bladder, brain, prostate, NHL, leukemias, laryngeal, soft tissue cancers, and so on; as opposed to women who had a drastic drop between the incidence of breast cancer and all other cancers. This observation is even more critical for making the decision of where to focus cancer screening efforts especially among women in a resource-constrained setting like Iraq. The IARC’s GLOBOCAN 2018 cancer incidence report had shown that the top five cancers in Iraq were breast, lung, leukemias, bladder, and colorectal cancers; which appears to be at odds with what was found in this study which had recorded breast, lung, brain, bladder, and NHL as being the top five.^2^ It is possible that being a single site study, these findings may be skewed, and a national study may be more relevant to comparing with the findings in the GLOBOCAN report. Nonetheless, several other researchers agree with the fact that breast cancer predominates and requires significant efforts at controlling it.^7,19,20^

Furthermore, majority of the patients presented with primary disease that were not a recurrence and not associated with distant metastases. This finding may again be as a result of the increasing awareness of cancers and improved cancer screening programs leading to early disease detection and treatment. This is also buttressed by the fact that a large proportion of the patients present with Grade I (11.31%) and II (21.35%) disease, suggesting timely presentation at cancer care centers for screening and necessary management. Even though there is sparse literature on the clinical profile of Iraqi cancer patients, findings from a breast cancer study by Mutar et al agrees with these results as it indicates that about 57.8% of the breast cancer patients studied presented in Grade 2 and another 12.9% presented in Grade 1.^7^ On histopathology however, the majority of the patients who had their disease staged were found to have Stage IV disease (10.96%, Table 3), followed by those in Stage IIIA (4.33%) and Stage IIB (4.08%). The presentation of the majority of patients with Stage IV disease which suggests that there is already metastatic disease is in line with the fact that as much as 20.88% of the patients were found to have had distant metastases upon evaluation. Comparing the stage and grade, it is important note that both do not necessarily need to agree as it is possible for a patient with Grade II disease to be in Stage IV of the illness.

The study conducted a deep dive into the pattern of presentation of the patients with breast cancer, as it was the most common (n=1164, 23.99%) of all the cancer types among the patients studied. The mean age varied from 49.35 years in 2016 to 50.61 years in 2018. This increase is not significant and agrees with findings from two recent studies that reported that the mean age of Iraqi breast cancer patients was 52.0 and 49.4 years.^7,19^ Other studies on women in countries such as Lebanon, Turkey, Bahrain, and Jordan within the middle eastern region have reported comparable findings.^21–23^ The disease was unilateral in the majority of cases with a comparable distribution between the left (33.25%) and right (31.96%) breasts. A recent study had reported similar findings, even though the right breast was affected in 50.6% of the patients as opposed to the 42.5% who had the left breast affected.^19^

Histopathologically, a large proportion of the breast cancer patients had ductal disease (82.90%), similar to the study by Karim et al which observed that as much as 83.17% of the study population had ductal cancers, followed by 5.94% tubular, 6.93% lobular, and other types.^19^ There is scarcity of data on the pathological profile of breast cancer patients in Iraq which makes it difficult to compare this study’s findings. However, with the relatively better prognosis of patients with ductal disease, especially when it is still at the ductal carcinoma in-situ stage, there may be a good chance for survival of the majority of breast cancer patients in the country. This again points at the need for aggressive screening, early treatment and follow up of breast cancer cases. Also, a sizeable proportion of the breast cancer patients expressed the three critical receptors influencing the prognosis of their disease. At least, 133 patients (29.75%) had amplified Her2 receptors, 323 patients (71.62%) were PR positive, and 347 (76.43%) were ER positive. This finding is crucial in helping clinicians determine the best treatment approach. It also suggests that another sizeable proportion of the patients might have been triple-negative (that is, not expressing any of these receptors on affected cells) which worsens their prognosis. Previous studies have indicated that up to 15.6% of the breast cancer population are triple negative, which may explain in part the high mortality rates in breast cancer patients.^24^

Overall, the findings in this study are vital to efforts by government, cancer care professionals, and advocates to be more aggressive in ensuring that middle-aged individuals who are more at risk of the various types of cancers are screened as a way of ensuring early detection and ultimately, control of the disease. The study has also shed more light on the demographic, clinical, and histopathological profile of cancers in Middle Euphrates area, which to some extent, reflects the country’s cancer epidemiological profile. One thing that is clear from available evidence is that the apparent rise in cancer incidence in this area of Iraq is due to the effect of a pack of factors rather than a single cause.

However, the study suffers from a few limitations, especially with regards to the fact that this was a single site study which limits generalizability and the inadequate collection of variables on the histopathological, genetic, and molecular profiles of cancer patients seen. The combination of the data in this study with molecular/genetic and survival data would have provided a more robust outlook on the cancer landscape in Middle Euphrates area, and more importantly, would have been helpful in developing a predictive artificial intelligence model for predicting who is likely to have cancer and what the potential outcomes will be. At a time like this when the world is shifting to more individualized care for cancers, the lack of detailed demographic, clinical, histopathological, and molecular/genetic information about cancer cases in the national cancer registry will only lead to poor cancer care outcomes, worse cancer indices, and waste of resources. It is therefore important that additional large-scale studies be carried out in the future to provide detailed information which can serve as input into the design of an AI model focused on streamlining and individualizing cancer care. Also, cancer registries and oncology databases need to be improved upon to capture all relevant details about patients with cancers.

## Conclusion

This study demonstrated the increasing burden of cancers in Iraq over time, and the predisposition of women to be more at risk of cancers, especially breast cancer. The findings here have vital implications toward reducing the incidence of breast cancers and other cancers by targeting middle-aged women and men with screening programs and supporting advocacy for early disease detection and care. Also, there is a need for the optimization of cancer registries and databases to capture detailed information on cancer patients which will be crucial towards the development of AI models that will help individualize and improve the outcomes of cancer care in Iraq.

**No additional data available for this study.**

## Data Availability

The study utilized a retrospective descriptive study design

